# Neural networks for dengue forecasting: a systematic review

**DOI:** 10.1101/2025.10.14.25338016

**Authors:** L. Lober, F. A. Rodrigues, K. I. O. Roster

**Affiliations:** Departamento de Matemática Aplicada e Estatística, Instituto de Ciências Matemáticas e de Computação, Universidade de São Paulo—Campus de São Carlos, Caixa Postal 668, 13560-970 São Carlos, São Paulo, Brazil; Harvard T. H. Chan School of Public Health, Boston, MA

**Keywords:** Neural networks, dengue, forecasting, machine learning, systematic review

## Abstract

**Background:** Early dengue forecasts are an important tool for disease mitigation, and neural networks (NNs) are powerful predictive models that have contributed to many areas of public health. We reviewed NN applications in the dengue forecasting literature to inform model design for future studies.

**Methods:** Following PRISMA guidelines, we systematically searched Web of Science/Knowledge, Scopus, PubMed, and Science Direct up to September 2025 for studies using NNs to forecast dengue in human populations, retrieving 628 records. After removing duplicates, four inclusion and two exclusion criteria were applied: articles had to be in English, implement NN or deep learning techniques, predict dengue fever incidence or risk, and clearly state data sources, processing steps, network parameters, and architectures.

**Results:** Sixty two papers were included and summarized by relative performance of neural networks and comparator models, architectures and hyper-parameters, choices of input features, geographic spread, and model transparency. Most studies (35 out of 62) implemented shallow feed-forward neural networks, using historical dengue incidence and climate variables as the most common input features, with 43 appearances. Target variables were dengue cases, severity and hotspots, and risk of outbreak, with model selection and evaluation approach being non uniform. Most studies (25) used granular observations at the city level or its subdivisions, while also commonly employing data from weekly measurements (35 out of 62). Performance of neural networks relative to comparators, such as tree-based supervised models, varied across study contexts. We found that 63% of studies included such models in their baseline; of these, approximately half found neural networks to outperform the baseline, although most do not include statistical comparisons.

**Conclusions:** The studies suggest that neural networks, although useful in specific scenarios, are not consistently found to be superior to simpler predictive models, especially tree-based machine learning methods.

## 1 Introduction

Dengue fever is a mosquito-borne disease with a significant global burden. More than 100 countries are at risk of infection [**1**], with a substantial increase registered in the last five years and spread to new geographic areas, such as Europe and eastern Mediterranean regions [**2**]. Dengue fever presents as a flu-like illness with symptoms ranging from mild to severe, with potentially life-threatening complications arising from severe dengue. The virus occurs in four serotypes, meaning up to four infections are possible for each person over their lifetime, and no specific treatment currently exists. Even with the availability of vaccines in many countries, dengue mitigation relies on vector control and early warnings to public health agencies [**1**, **3**, **4**]. To that end, predictive models can help efficiently allocate public health resources to combat dengue, with the goal of reducing the overall disease burden.

The literature on dengue forecasting is multi-disciplinary, uniting expertise from areas such as epidemiology, environmental science, computer science, and mathematics. Modeling frameworks include both theoretical and data-driven approaches, spanning dynamic transmission models, statistical time series forecasting, and machine learning methods. Dengue incidence is associated with many different risk factors, which may be included as variables in predictive models. These include climate conditions such as rainfall, extreme weather events, and temperature [**5**, **6**, **7**], land use [**8**], and poverty [**9**]. These factors, and others related to dengue, may have nonlinear, context-specific, and time-varying effects on disease incidence, which poses a challenge to disease modeling.

Machine learning models benefit from the increasing data availability on risk factors on the spreading of diseases, and offer non-parametric approaches that require less detailed knowledge of the disease and its context [**10**]. Neural networks (NNs) are a subset of these algorithms, which have made significant contributions to medicine and public health, including applications such as medical image analysis for disease diagnosis [**11**], identifying abnormalities in signals such as electrocardiographs (ECG) [**12**], and optimizing decisions of health care providers, hospitals, and policy-makers [**13**]. Neural networks are also used to forecast diseases, including malaria [**14**], influenza [**15**], and COVID-19 [**16**].

The aim of this review is to describe the technical decisions made in the literature on dengue forecasting with neural networks, including architecture selection, hyper-parameter tuning, choice of input variables. We also aim to assess the relative performance of different neural network architectures and comparator models, such as other machine learning techniques.

## 2 Methodology

To achieve the aforementioned objective, this review followed the Preferred Reporting Items for Systematic Reviews and Meta-Analyses (PRISMA) statement guidelines [**17**]. The PRISMA flow chart for this study is represented in Figure 1. A systematic search was conducted using Web of Science/Knowledge, Scopus (abstract, title, keywords), PubMed, and Science Direct (abstract, title) databases. References of papers appearing in the search results were also examined for relevant works. The searches were conducted on September 10, 2024, and used the following search string:

> “(deep learning OR neural network) AND dengue”

**Figure 1:**
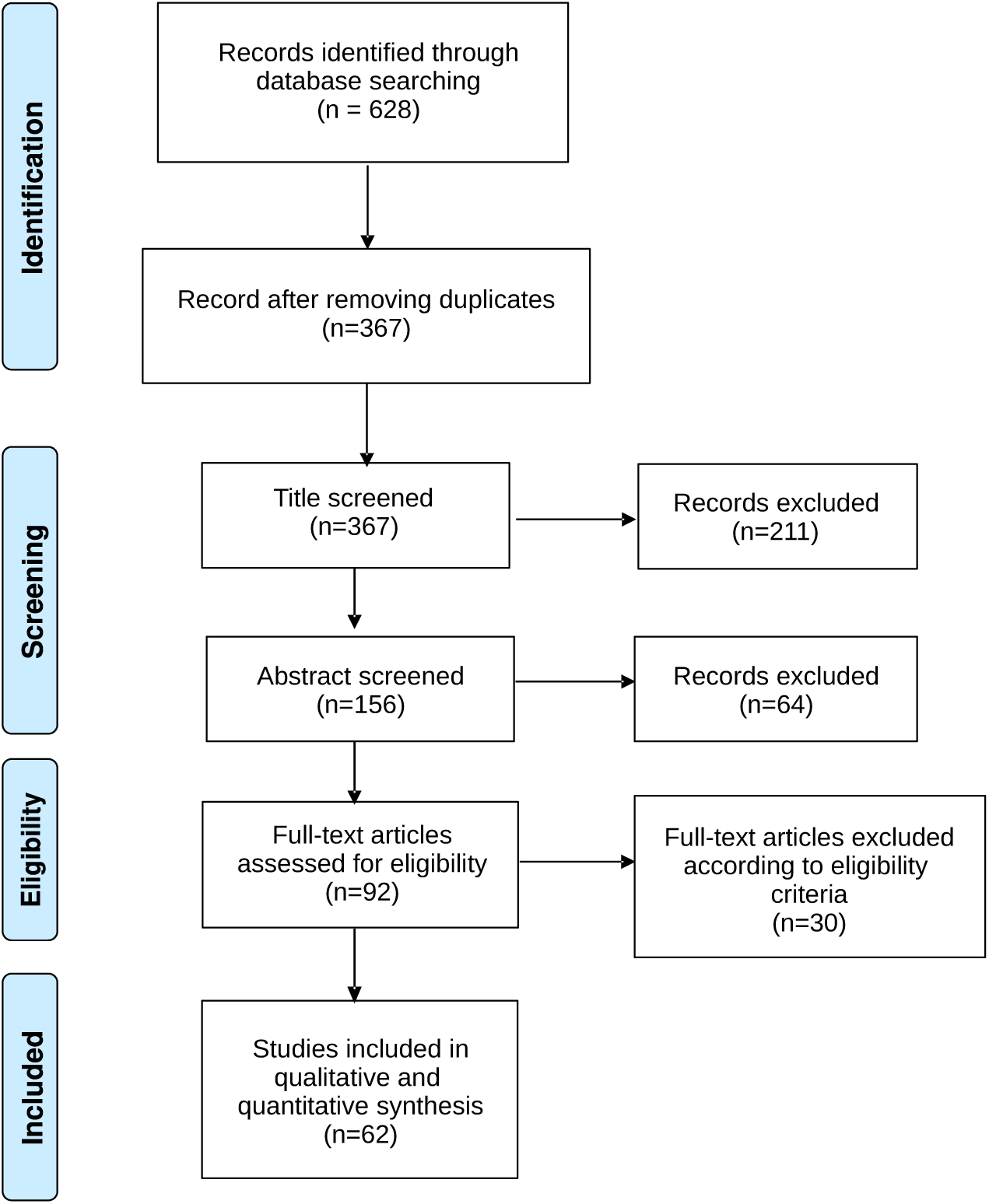
PRISMA [17] flow chart for this study.

All data collected from the database search were compiled in a spreadsheet. The papers were then examined for inclusion in the review in three stages: by title, by abstract, and finally by full text. In each phase, the following inclusion criteria were applied conservatively. If the information in the title and/or abstract was inconclusive, the paper was included in the full-text review. These inclusion criteria were as follows:

- Studies must implement a neural network or deep learning technique, either as the main method or as a comparator model. Reviews of the literature on neural networks for dengue forecasting were also considered;
- Studies must predict dengue fever incidence or risk. There were no restrictions on the type of target variable used. For example, the number of dengue cases, dengue incidence rate, or a binary dengue risk variable were all accepted as target features;
- Studies must examine dengue in a human population, excluding studies that forecast vector densities or similarly motivated entomological research. Studies focusing on disease diagnosis, animal hosts, or the location of vectors without relation to dengue incidence were excluded;
- As the search query is in English, only English language articles were identified and included in the review.

Alongside these criteria, two other standards were used when screening the full texts of selected papers:

- Data sources and processing steps must be clearly stated;
- Methodology must clearly state neural network parameters and architecture.

We also excluded papers that did not allow institutional access to the authors for reviewing, which was the case for three studies that fulfilled the eligibility criteria up to the full-text screening stage. The results of each stage of the screening process are listed in the PRISMA flowchart in Figure 1.

The following information was extracted from each study: open access status, access to data, source code status, publication medium, year of publication, country, geographic level of aggregation, target variable, prediction period, input data, variable details, data level of aggregation, train/validation/test split, NN model type, model categories, architecture details, comparator models, model evaluation metric, results (descriptive from text), and best model listed.

No automation tools besides the downloading of databases and data aggregation for figure plots were used in this review. The Python notebook used for analysis and visualization is available in [**18**]. All authors contributed to the assessment of the quality of extracted data independently.

## 3 Results

### 3.1 Data Sources

Figure 2 shows meteorological features as the most common additional data source, being featured in 43 of the 61 papers included in this study. This data comprises temperature, humidity, wind speed, solar radiation, pressure, rainfall, sea surface temperature measurements, and El Niño data.

**Figure 2:**
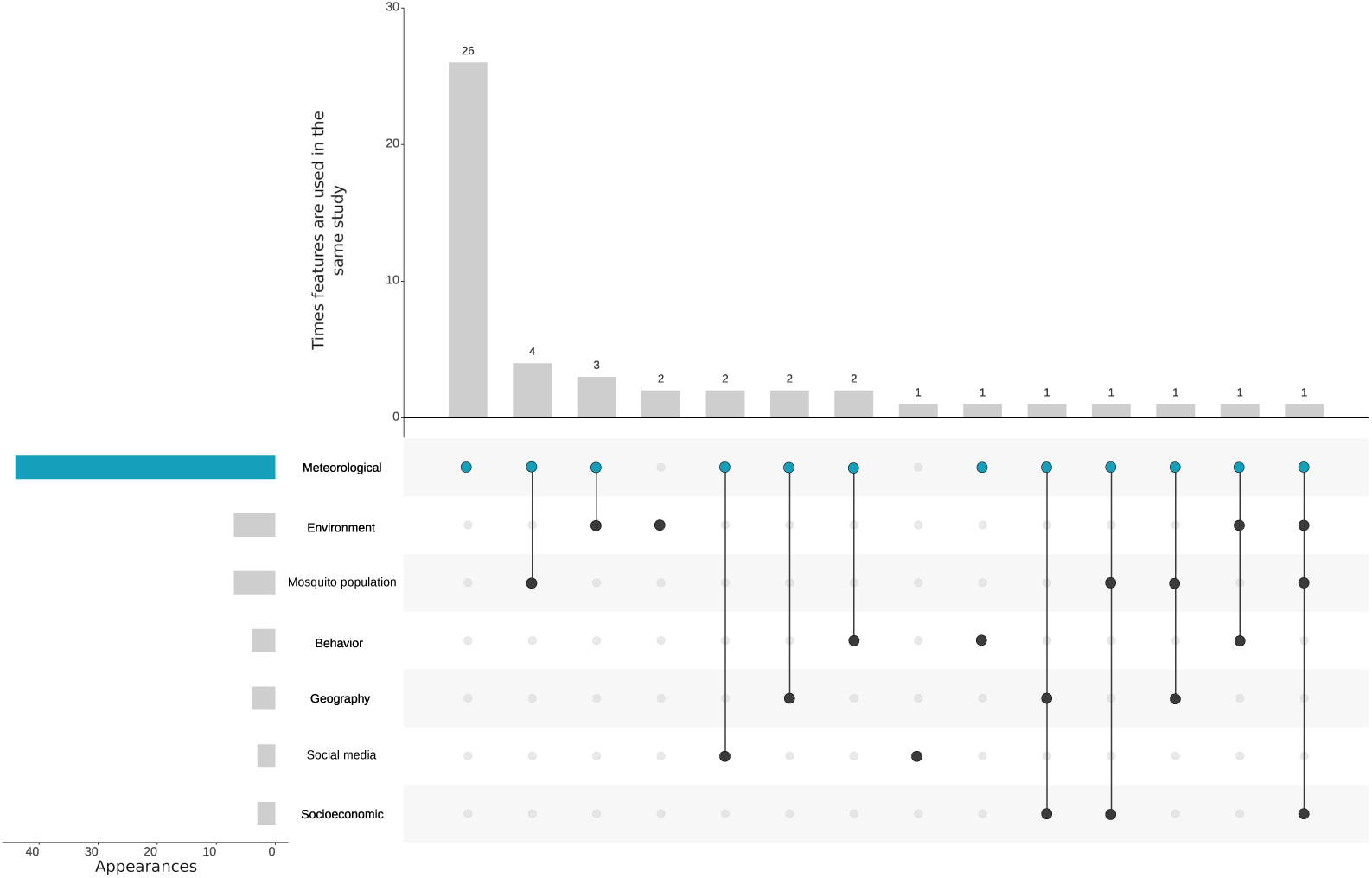
Input data. Frequency with which studies included predictor variables and their combinations as input data for prediction. The top bars measure the number of papers that use each combination of variables shown as connected dots in the central plot. The left horizontal bars show the frequency with which each individual category is included in the selected studies. ‘Meteorological’ data is present in most (69%) studies.

Out of all papers that did not include meteorological data, only four [**20**, **21**, **22**, **23**] added extra features to train the neural networks beyond the epidemiologic information on dengue fever.

In Anderson et al. [**20**, **21**], both studies combined environmental data from satellite and street-level imagery with epidemiological data for their forecasts; Livelo and Cheng [**22**] used social media data to complement their models. Araújo et al. [**23**] employed data on bus lines to understand how mobility restrictions in the pandemic years impacted dengue transmission in urban areas.

The second most frequent data source used for dengue fever forecasting was environmental data, found both as satellite images [**24**], vegetation indexes [**25**, **26**, **27**, **28**], and street imaging from the Google StreetView program [**20**, **21**].

Following closely the number of uses of environment information, data on mosquitoes for forecasting dengue included mosquito vector density [**27**] and density per person in a given month [**29**], larvae free factor [**30**], biting rate [**31**], entomological data in the form of raw number of *Aedes aegypti* and *Aedes albopictus* mosquitoes [**32**], and also the infection rate of larvae, male, and female mosquitoes [**33**]. Moreover, Hu et al. [**28**] used species distribution models for dengue-transmitting mosquitoes and *Aedes aegypti* temperature suitability.

Other, rarely occurring features found across the studies were human behavior, social media, geographic, and socioeconomic data, which includes:

- **Behavior:** Number of holidays in a given week, used as a boolean variable by Bergero et al. [**34**], human mobility through call data records (CDR) processing [**25**], bus routes [**23**], and time spent in transit stations from Google Regional Mobility Reports [**35**];
- **Social media:** Short messages from X (formerly Twitter) containing dengue-related information, selected through preprocessing methodologies to ascertain the relevance of the social media data to dengue circulation [**36**, **22**, **37**];
- **Geography:** Anno et al. [**38**] included this information from data on the place of residence of infected individuals, labeled as township or village. We also listed population density from Kesorn et al. and Das et al. [**33**, **39**] considering land use, explicitly used in Francisco et al. [**40**], to dengue forecast.
- **Socioeconomic data:** Dong et al. [**32**] incorporated multiple socioeconomic variables, including literacy rates, healthcare access, and sanitation infrastructure, which included houses with dirt floors, toilet facility, water pipelines, sewage system, and electricity.

Epidemiological data, e.g. case counts or incidence rates, are necessarily included in all studies due to the nature of the prediction problem.

Studies focused both on macro regions or smaller divisions of a given area for their forecasts, covering forecasts at district-, city-, province-, and country-level (Figure 3). “Province” denotes first-level administrative subdivisions (e.g., states), whereas “district” refers to intra-city subdivisions, which include both neighborhoods and the health center coverage area within a city. In Figure 4, Brazil was the most frequent country in our sample, with 12 papers using Brazilian data in their analyses, followed by Indonesia (8 papers) and India (5 papers).

**Figure 3:**
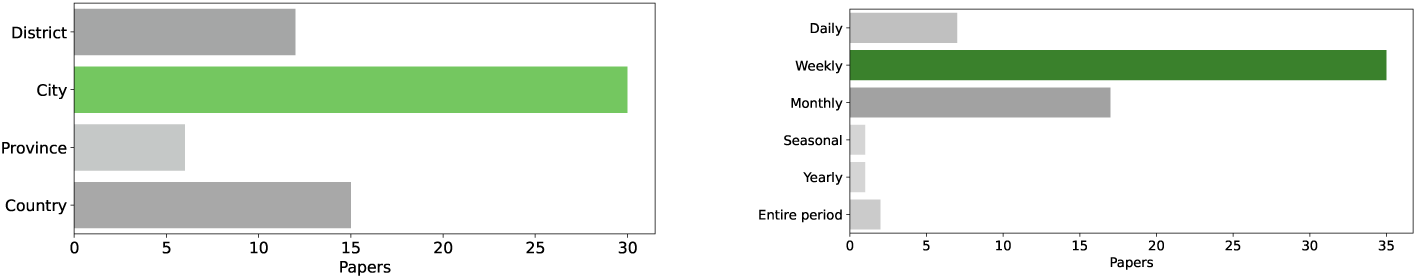
Geographic and temporal distribution of forecasting scale, with green bars highlighting the most frequent category. The left panel shows the frequency with which studies predict dengue cases at the city, district, province, and country level. The right panel shows the frequency with which studies employ data and predict target variables at daily, weekly, monthly, seasonal, and yearly levels. The “entire period” of dengue data accounts for the total number of cases in a certain period, and was used by studies that predicted dengue fever risk in specific regions [**19**, **20**].

**Figure 4:**
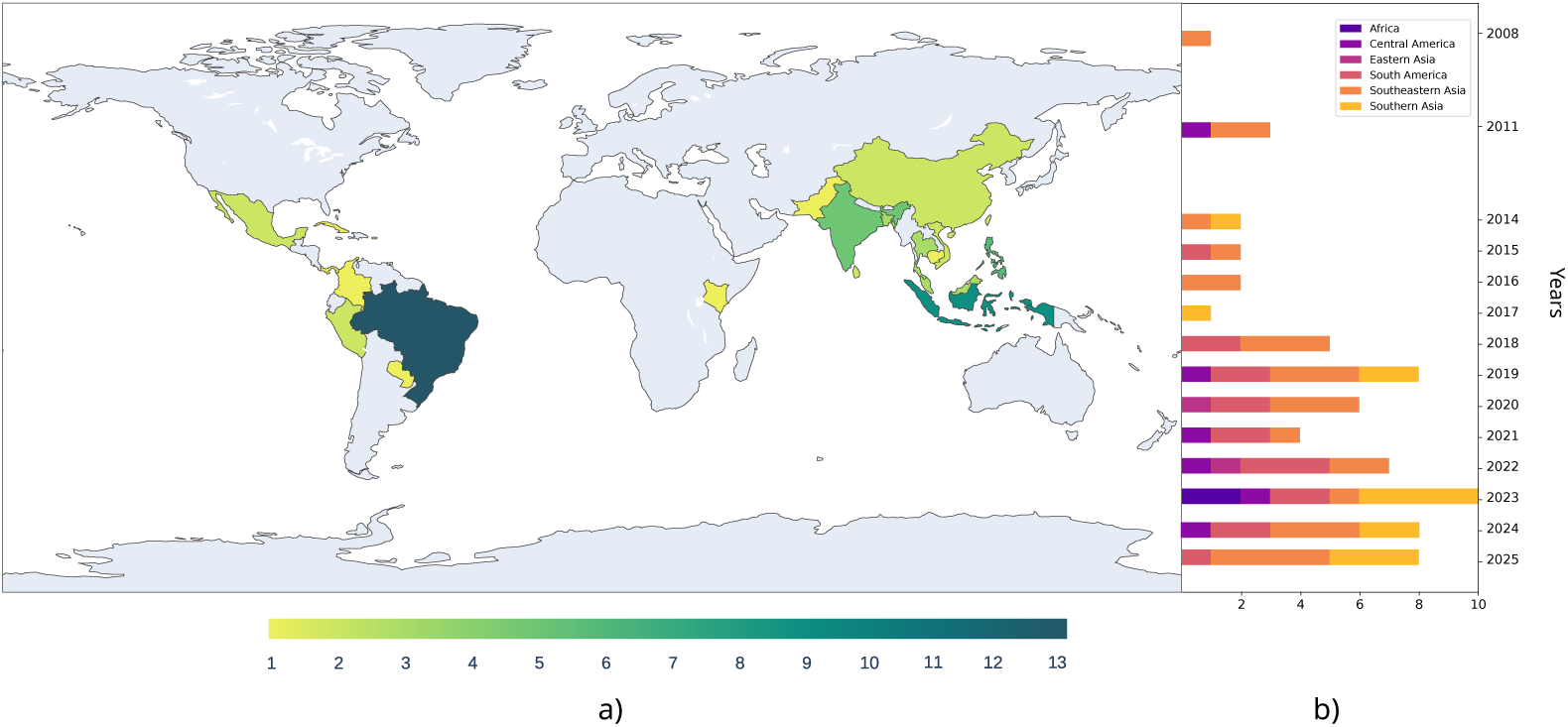
Geographic coverage. a) Number of studies in each country from 2008 to 2025 and b) number of studies in different geographic regions, by year of publication.

We also analyzed the accessibility criteria of papers, verifying data and code availability in the full text, public accessibility of the paper, and publication venue, with results condensed in Figure 5. We split papers not published as “open access” in two categories, to understand whether the authors made the full text available as a pre-print before publication. With that in mind, 71% of the selected studies allow for free access to their methodology and results.

**Figure 5:**
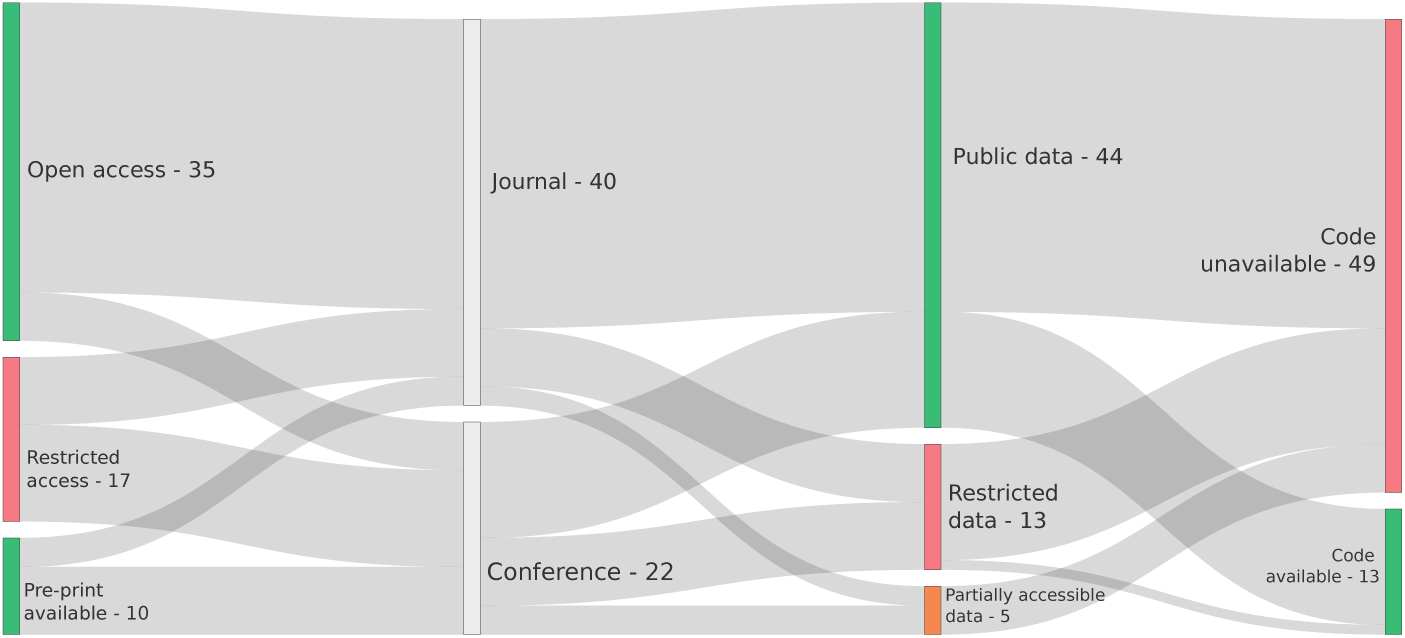
Accessibility of studies. The vertical bars show the frequency of open and restricted access, publication venue, data availability, and source code availability of the selected studies. The gray flows indicate how frequently combinations of characteristics appear together.

As for the data being readily available from the listed references, some cases included sensitive data on a population (such as call data records), and as such had only partially published data sources alongside the papers, being then labeled as “partial access to data” in Figure 5. As for papers with restrictions on the data sources, most of them either referenced sources no longer available, or that required formal data-use authorization from host institutions.

Lastly, only one fifth of all papers were published alongside their source code, with almost all that do also coming from open access journals. From those, the policies of most of those journals were to encourage authors to also make their code available, being only a requirement at *Nature Communications: Medicine* with Dong et al. [**32**], *PLOS ONE* with Baquero et al. [**41**], and *PLOS Neglected Tropical Diseases* with Araújo et al. and Francisco et al. [**23**, **40**].

### 3.2 Architectures

We reviewed the implemented architectures of neural network algorithms from the selected studies, distinguishing recurrent neural networks (RNNs), convolutional neural networks (CNNs), feed-forward artificial neural networks (ANNs), and other architectures. We also analyzed hyper-parameters and other modeling choices, such as network size (hidden layers and units), regularization (dropout), and learning rate. In total, shallow NNs, defined as architectures with up to one hidden layer, were the most prevalent (40 out of the 95 implemented models), followed by deeper architectures (36 RNNs and its subtypes, 8 CNNs) and 12 uniquely appearing algorithms (Appendix A). A sum of appearances of these architectures over the years for the select papers is shown in Figure 6.

**Figure 6:**
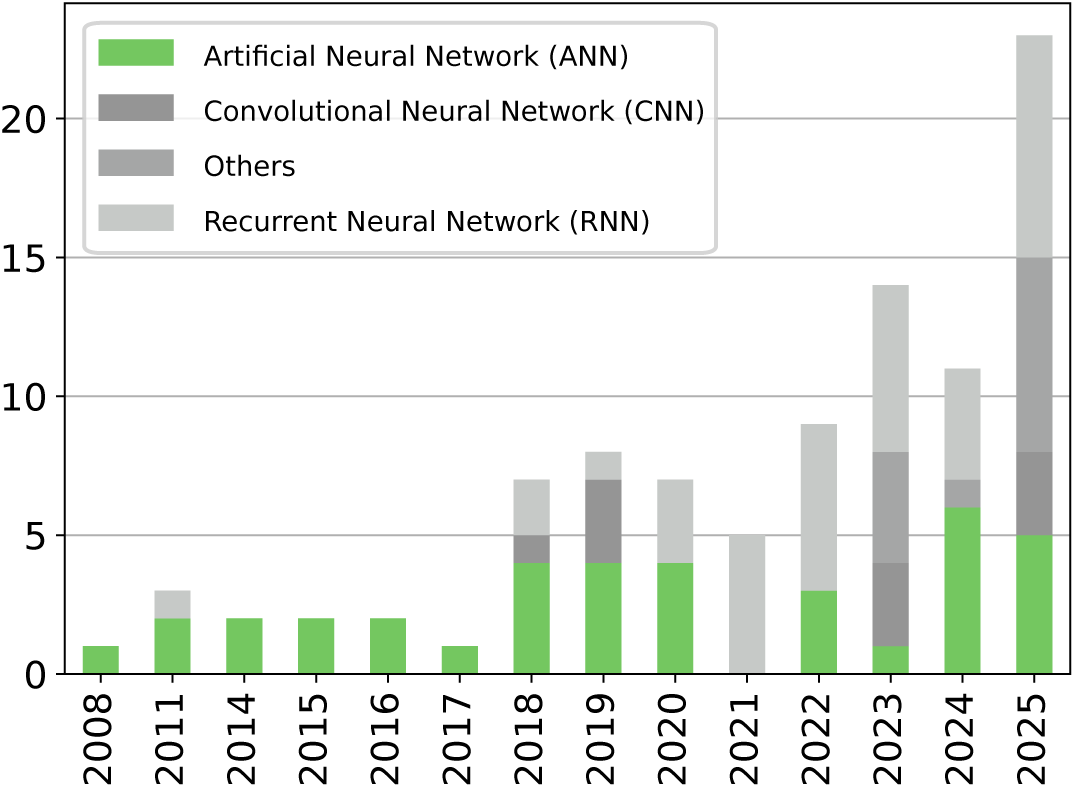
Types of neural network architectures over time. The green bars indicate the most commonly used architecture type.

Most studies implementing ANNs utilized standard fully connected structures, with a few exceptions being given by the NNAR in [**42**, **43**, **44**, **45**, **46**], an autoregressive neural network algorithm that relies on statistical criteria for parameter tuning and data processing; and the RBFNN in Muhammad et al. [**47**], which uses radial basis functions as activation functions in the output layer. One study [**48**] uniquely implemented a Fuzzy system in order to use clustering for dengue forecast, while also comparing their method to other clustering algorithms.

Two studies featuring CNNs [**38**, **24**] implemented a U-Net, which is a convolutional neural network architecture widely used for image segmentation [**49**], to combine satellite images and epidemiologic data to forecast dengue infections in a given region. All others employed the usual convolutional layer stacking of CNNs, with varying number of layers and parameters (see Table **??**). Among RNN architectures, Long Short-Term Memory (LSTM) networks were the most widely applied, appearing in 20 studies. Other less frequent architectures are given by the gated recurrent units (GRU) in [**50**, **51**, **52**, **22**] and the BlockRNN [**46**]. A Hopfield network is used in García-Garaluz et al. [**53**] to infer the reproduction number *R*_0_ in a Susceptible-Infectious-Recovered (SIR) compartmental dynamic transmission model, integrating the resulting dynamic system to evaluate performance against observed epidemiological time series. Other studies [**24**, **31**] also employed neural networks and compartmental models for forecasting. All mentions of optimizer usage were found in this category, with six studies opting for the Adam optimizer [**50**, **52**, **54**, **26**, **55**, **41**, **56**, **39**], and one each for Adamax [**55**] and RMSProp [**22**].

**Table 1:** Hyper-parameter values by neural network (NN) architecture type. Appearances refers to the number of papers using a neural network architecture that also report a given parameter. As some models define the size of layers or number of hidden units according to the number of lags, namely the number of steps delaying a time series, this information is included here for clarity.

| Architecture category | Parameter | Min. value | Max. value | Median | Standard deviation | Appearances |
|---|---|---|---|---|---|---|
| **Artificial NN** | Epochs | 85 | 8000 | 500 | 2679 | 11 |
|  | Hidden layers | 1 | 100 | 1 | 20 | 30 |
|  | Hidden units | 1 | 256 | 6 | 53 | 26 |
|  | Lags | 0 | 17 | 4 | 5 | 10 |
|  | Learning rate | 1E-5 | 9E-1 | 3E-1 | 0.3 | 4 |
| **Convolutional NN** | Batch size | 32 | 100 | 66 | 48.1 | 2 |
|  | Dropout | 0 | 0.5 | 0.25 | 0.2 | 3 |
|  | Epochs | 100 | 1000 | 550 | 567.4 | 4 |
|  | Total layers | 2 | 161 | 27 | 70.0 | 4 |
| **Recurrent NN** | Batch size | 10 | 204 | 32 | 57 | 9 |
|  | Dropout | 0 | 0.8 | 0.25 | 0.22 | 14 |
|  | Epochs | 10 | 8000 | 300 | 1651 | 14 |
|  | Hidden layers | 1 | 64 | 3 | 15 | 15 |
|  | Hidden units | 2 | 1266 | 32 | 344 | 16 |
|  | Lags | 1 | 16 | 9 | 4 | 5 |
|  | Learning rate | 5E-6 | 1E-2 | 2E-3 | 4E-3 | 9 |

From the 13 papers that discuss activation functions, it was found that Rectifier Linear Units (ReLU) was the most commonly used activation function, found in six studies [**36**, **51**, **38**, **57**, **41**, **21**], followed by the sigmoid activation function [**58**, **59**, **29**, **30**]. Studies also used the logistic sigmoid [**60**], logistic [**61**], hyperbolic tangent (tanh) [**36**], linear [**29**] activation functions, with the last two used in a layer before the dense layer.

Uniquely mentioned parameters are the Akaike information criterion (AIC) in [**46**], used to select the size of the input layer; the spread factor *σ* for a particle swarm optimization (PSO) algorithm found on Anggraeni et al. [**27**]; momentum on Anggraeni et al. [**58**], and Hasanah et al. [**48**] utilized fuzzy IF-THEN rules to create input-output pairs for an ANN+fuzzy layer hybrid. Pre-trained embeddings are also used in Livelo et al. [**22**].

### 3.3 Model selection and evaluation

Most studies implemented multiple algorithms, comparing different neural network architectures or machine learning models, as shown in Figure 7. ARIMA, being by far the most frequent statistical-based comparator model, was set apart from other similar models, while supervised models were split into tree-based methods, such as XGBoost and Random Forests, and other algorithms. Figure 10 shows the variety of NN and comparator models found in this review, alongside their number of appearances.

**Figure 7:**
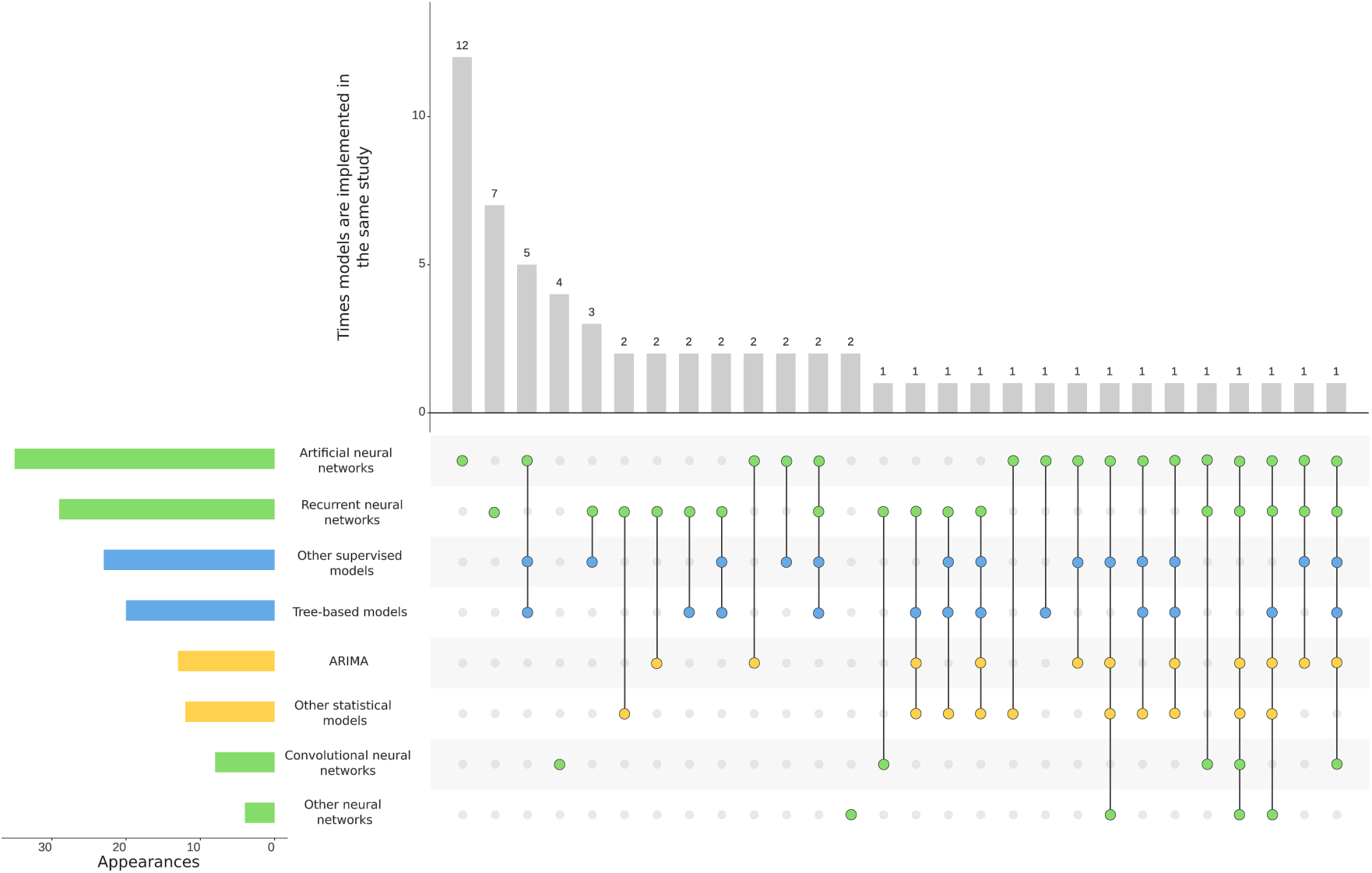
Frequency of comparator models. The horizontal bars show model frequency in the selected studies, with colors grouping structural similarity of these models (green: neural networks, blue: supervised models and yellow: statistical models). Vertical bars show the frequency with which combinations of models were used in a given study.

Out of the 10 studies that used CNNs, half of them do not employ another comparator model, which is also the highest rate out of all neural network categories (11 out of 34 for ANNs and 6 out of 28 for RNNs).

When studies did include comparator models, ANNs tended to feature statistical models more often (14 out of 34 entries), with ARIMA being particularly prominent in this case, with 9 papers, while other papers used supervised models for comparisons. RNNs have the opposite distribution, with studies having used supervised models more often and only 10 applying statistical models. Finally, two studies used CNNs as baseline comparators against novel proposed models: one for an original wavelet neural network architecture [**46**], and another using multiple CNN and RNN architectures [**51**]. Others compared the CNNs to other neural networks [**62**], statistical models and tree-based algorithms [**63**], and an ensemble of all three model types [**37**].

Notably, 21 out of all studies included lags, which are delays in a time series set according to a window size defined by the user. They were used in these studies for both epidemic records and the auxiliary variables. Moreover, the 11 studies that also use any type of statistical model as comparators, such as the various autoregressive algorithms shown in Figure 10, favor employing lagged time series to the modeling process, with 65% of them having this type of feature in the training dataset.

Within the criteria of study selection, there was significant variation in the studies’ specifications, including the formulation of the target feature. Most studies (88%) predicted the future number of dengue cases based on historical time series. Others made forecasts on the risk of outbreak or dengue severity and its hotspots in a given region. The metrics found throughout this review were grouped accordingly in Figure 8.

**Figure 8:**
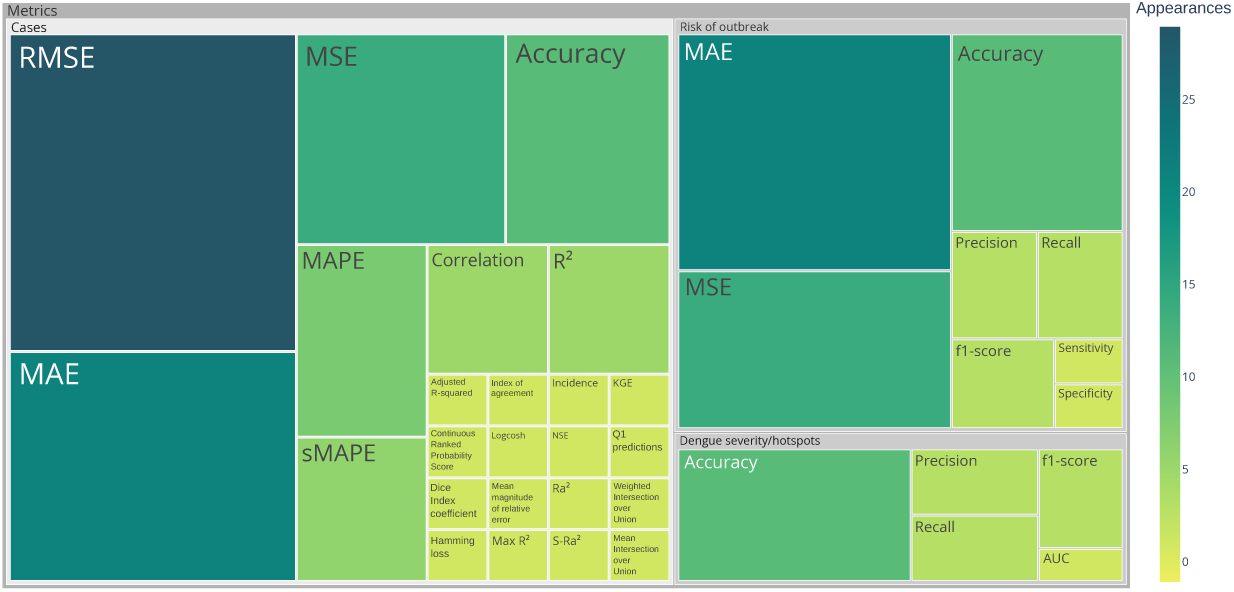
All performance metrics found in the select papers, grouped by target variable: dengue cases forecast, risk of outbreak and dengue severity or hotspots.

Another important distinction between studies is that the forecast window varies depending on the predictive objective and not all studies reported prediction windows: most of the studies (44%) made predictions for the whole testing set, which varies in size depending on the data split chosen by authors. When reported, forecast horizons ranged from 1 to 12 weeks ahead, with the most common forecasting windows employed by these studies ranging from one up to four weeks. Lu et al. [**31**] included multiple forecast windows, which were treated separately when accounting for the data length displayed in Figure 9.

**Figure 9:**
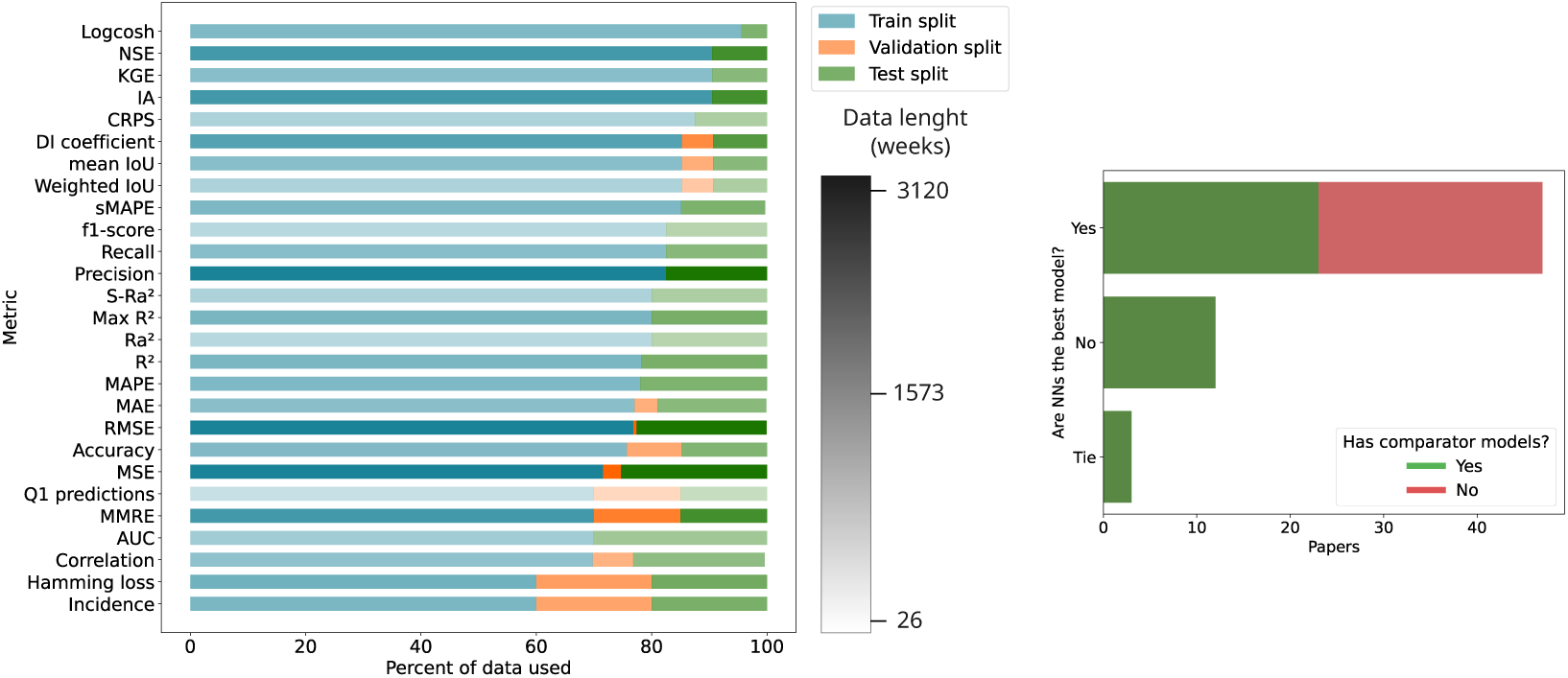
Model evaluation. The left panel shows the average observed splits for each metric, with color gradients indicating the total length of data employed. The right panel shows whether neural networks were stated by authors to be the best performing method, and whether the study included comparator models.

**Figure 10:**
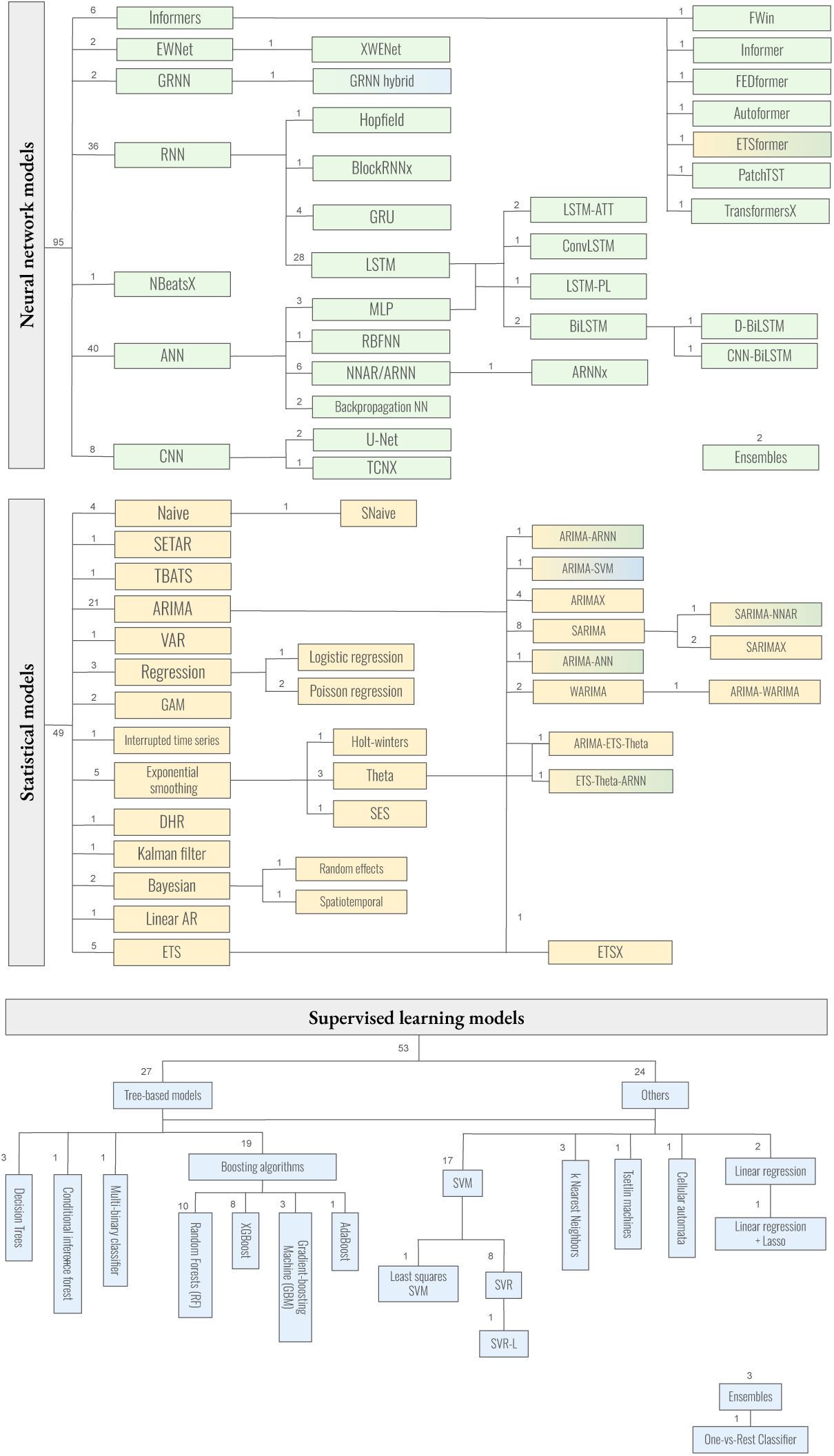
All models found throughout the screening process and included in this review, grouped by structural similarities.

From Figure 8, the most common metrics by type of forecast are: Root Mean Squared Error (RMSE) for forecasting dengue fever case counts, Mean Absolute Error (MAE) for risk of outbreak, and accuracy for detecting dengue severity, or hotspots, in a given area. More than half (58%) of these studies also employed more than one evaluation metric. For niche metrics, only three studies [**64**, **65**, **23**] do not include more frequently used ones when evaluating models through multiple measurements.

It was also found that there was no standardized protocol for acquiring and preprocessing datasets to train neural networks for dengue forecast, as shown by the different choices in comparison to the type of metric used. Average time series length, in weeks, was found to be 457.83*±*558.00 with *σ*^2^ = 209607.13. Even when considering the same geographic scale, it was rare that studies shared the same granularity, target location and time series length choices. Cross-validation is also used sparingly, and used specifically in [**66**, **61**, **67**, **53**, **29**, **65**, **68**, **63**, **31**]. Notably, 23 out of the 47 papers that do affirm that neural networks are the best approach to dengue forecasting for a given location only discussed a single architecture with no comparator models, at most comparing their results to other similar applications from the literature, as shown in the right panel of Figure 9.

## 4 Discussion

We reviewed the literature on dengue forecasting using neural networks to analyze choices in model construction and provided a practical guide for researchers aiming to implement neural networks in their own forecasting work. Nearly half (49%), or 23 out of 47 studies, included in this systematic review reported that their proposed neural network architectures outperformed other machine learning or statistical approaches, suggesting that neural networks may be promising candidate models for dengue forecasting. However, given positive publication bias and limited comparator models in some studies, this result suggests that neural networks are not inherently the most superior model and other machine learning and statistical approaches, including ARIMA and tree-based methods often out-perform neural networks, while also offering greater interpretability. As neural network architectures are advancing, their success rate may improve, but the current state of the literature suggests that neural networks should be considered as only one of many possible algorithms for dengue forecasting problems.

Though many risk factors of dengue have been identified, most neural network models in this review limited inputs to meteorological and epidemiological data. Alternative sources of predictor variables leveraged by studies in this review include data from X (formerly Twitter) [**36**, **22**, **37**], Baidu search data [**10**], Google Street View and satellite images including derived features [**21**, **25**, **24**, **28**, **40**], mobile phone records [**25**], and mobility data [**23**, **35**]. Future studies may evaluate the value of alternative predictors, especially as neural networks can be implemented to handle high-dimensional problems well [**69**].

While datasets are expanding, there are still limits to dengue surveillance, including high asymptomatic rates and limits to diagnostic capacity [**1**]. One study in this review [**66**] aimed to address data scarcity using transfer learning: the authors trained a model on a city with high dengue incidence and later used it to predict the disease in lower-incidence geographies. This was the only study in this review that used transfer learning outside the context of pre-trained CNNs. This may present another avenue for further research, especially in locations where data is scarce, or to tackle under-reporting of dengue cases. Another paper aimed to improve dengue data by addressing the common problem of seasonal fluctuations in case counts, especially at fine temporal and spatial resolution, i.e., zero-inflation, which may lead to inaccurate predictions and performance evaluation [**40**]. The authors demonstrated that sparse data interfered with machine learning model accuracy. More research in this space can provide an avenue for a reliable use of data with higher granularity in the future.

There is increasing interest in improving explainability of machine learning models [**70**], which is also reflected in this review. For example, SHapley Additive exPlanations (SHAP) were used to locate climatic drivers of dengue outbreaks [**56**], although the usage of such tools is still rare and only found for these two studies in dengue forecasting. Statistical evaluation of NN performance was found only on [**28**, **35**], with other studies only including comparator models such as ARIMA as a statistical basis and only one study [**27**] performing systematic comparisons of their proposed approach to more than one dataset.

One study [**28**] used an ANN to provide global and local projections of dengue distribution and infection considering changes in temporal patterns of climate and socioeconomic data. Changes in climate are expected to greatly impact the global burden of infectious diseases, including dengue [**71**]. While much of the reviewed literature focuses on prediction in unchanged conditions, more work is needed to anticipate dengue outbreaks in places with profound structural changes in climate, urbanization, and other factors, which may disrupt historic patterns of dengue seasonality, the scale and scope of outbreaks, and endemicity. These models may prove critical to anticipating and controlling dengue outbreaks under variable conditions.

We were not able to conduct a meta-analysis of the performance of neural networks in the selected studies, due to limited overlap in datasets and geographies, variations in model architectures, including the train-test split, and lack of open access and incomplete reporting in model implementation and methods, which can include training time, inference time, and hardware requirements for the observed neural network architectures. Moreover, only a few studies [**25**, **29**, **60**, **80**] do include explicit comparison to other works in the literature – both for parameter selection and performance comparison. This limits our ability to generalize performance results of modeling choices in different contexts, such as epidemic vs. endemic regions of dengue. Future forecasting studies would benefit from comparing several different model types and architectures on multiple benchmark datasets to better understand how the proposed approach performs by their geographic context or data availability.

The field would also benefit from greater transparency and open source coding, as much of the code and data was restricted in the selected studies. One particular study proposed an online and modular platform to automate data retrieval and forecasting for dengue and other arboviruses of interest through several supervised approaches, including multiple possible neural network implementations, which can also be a valuable tool to guide health policies by enabling systematic benchmarking of these methods and datasets [**72**]. Efforts like these can help improve usability of and trust in advanced neural network models by governments to inform dengue control policies.

Neural networks are a promising and increasingly used tool in dengue forecasting. While much of the field still relies on simple architectures and datasets, several studies have pioneered the inclusion of diverse digital data streams, novel methodologies, such as transfer learning or satellite data. Prediction in the context of changing data generation mechanisms, for example due to climate change, is another promising new avenue of inquiry. However, more work is needed to compare model performance across geographies and settings and improve the transparency and accessibility of models for policymakers.

## Data Availability

The present study did not produce additional data, and all databases screened for this review are available online at PubMed, Scopus, Web of Science and Science Direct.

## Acknowledgments

Luiza Lober thanks the support given by Fundação de Amparo á Pesquisa do Estado de São Paulo (FAPESP) (grants number 2022/16065-3 and 2013/07375-0). Francisco A. Rodrigues acknowledges CNPq (grant 308162/2023-4) and FAPESP (grants 20/09835-1 and 13/07375-0) for the financial support given for this research. Kirstin Oliveira Roster gratefully acknowledges support from the São Paulo Research Foundation (FAPESP) (grant number 2019/26595-7).

## Declarations

This manuscript has been made available as a preprint at arXiv (arXiv:2106.12905) and medRxiv (10.1101/2025.10.14.25338016). No generative AI tools were used in the development of this study or writing of this manuscript. We declare that there are no competing interests of review authors.

## Notes

### Competing Interest Statement

The authors have declared no competing interest.

### Summary of Updates

Updated text to better clarify the scope of this systematic review and reflect PRISMA's guidelines and checklist.

## References

[1] World Health Organization (WHO). Dengue – Global Situation. [Accessed 25. Sep. 2025]. 2024 May 30. Available from: https://www.who.int/news-room/fact-sheets/detail/dengue-and-severe-dengue

[2] European Centre for Disease Prevention and Control. Dengue worldwide overview. [Accessed 26. Aug. 2025]. Available from: https://www.ecdc.europa.eu/en/dengue-monthly

[3] Takeda Pharmaceutical Company. Takeda and Biological E. Limited Collaborate to Accelerate Access to Dengue Vaccine in Endemic Areas. [Online; accessed 26. Aug. 2025]. 2025 Jun. Available from: https://www.takeda.com/newsroom/newsreleases/2024/collaboration-to-accelerate-access-to-dengue-vaccine

[4] Ranzani OT, Neto FL, Mareto LK, Brumatti TS, Oliveira RD de, Silva PV da, Santos ER dos, Agostini TLD, De Paula RAC, Dean NE, Ko AI, Cummings DAT, Andrews JR, Hitchings MDT, and Croda J. Effectiveness of the TAK-003 dengue vaccine in adolescents during the 2024 outbreak in São Paulo, Brazil: a test-negative, case–control study. Lancet Infect. Dis. 2025 Aug; 0. doi: 10.1016/S1473-3099(25)00382-2

[5] Chien LC and Yu HL. Impact of meteorological factors on the spatiotemporal patterns of dengue fever incidence. Environ. Int. 2014 Dec; 73:46–56. doi: 10.1016/j.envint.2014.06.018

[6] Thu HM, Aye KM, and Thein S. The effect of temperature and humidity on dengue virus propagation in *Aedes aegypti* mosquitos. Southeast Asian J. Trop. Med. Public Health 1998 Jun; 29:280–4. eprint: 9886113

[7] Fan J, Lin H, Wang C, Bai L, Yang S, Chu C, Yang W, and Liu Q. Identifying the high-risk areas and associated meteorological factors of dengue transmission in Guangdong Province, China from 2005 to 2011. Epidemiol. Infect. 2014 Mar; 142:634–43. doi: 10.1017/S0950268813001519

[8] Kilpatrick AM and Randolph SE. Drivers, dynamics, and control of emerging vector-borne zoonotic diseases. Lancet 2012 Dec; 380:1946–55. doi: 10.1016/S0140-6736(12)61151-9

[9] Mulligan K, Dixon J, Sinn CLJ, and Elliott SJ. Is dengue a disease of poverty? A systematic review. Pathogens and Global Health 2015 Feb

[10] Guo P, Liu T, Zhang Q, Wang L, Xiao J, Zhang Q, Luo G, Li Z, He J, Zhang Y, and Ma W. Developing a dengue forecast model using machine learning: A case study in China. PLoS Negl. Trop. Dis. 2017; 11:e0005973. doi: 10.1371/journal.pntd.0005973

[11] Mienye ID, Swart TG, Obaido G, Jordan M, and Ilono P. Deep Convolutional Neural Networks in Medical Image Analysis: A Review. Information 2025 Mar; 16:195. doi: 10.3390/info16030195

[12] Haglin JM, Jimenez G, and Eltorai AEM. Artificial neural networks in medicine. Health Technol. 2019 Jan; 9:1–6. doi: 10.1007/s12553-018-0244-4

[13] Shahid N, Rappon T, and Berta W. Applications of artificial neural networks in health care organizational decision-making: A scoping review. PLoS One 2019 Feb; 14:e0212356. doi: 10.1371/journal.pone.0212356

[14] Zinszer K, Verma AD, Charland K, Brewer TF, Brownstein JS, Sun Z, and Buckeridge DL. A scoping review of malaria forecasting: past work and future directions. BMJ Open 2012. doi: 10.1136/bmjopen-2012-001992.

[15] Alessa A and Faezipour M. A review of influenza detection and prediction through social networking sites. Theor. Biol. Med. Model. 2018 Dec; 15:1–27. doi: 10.1186/s12976-017-0074-5

[16] Bullock J, Luccioni A, Pham KH, Lam CSN, and Luengo-Oroz M. Mapping the landscape of Artificial Intelligence applications against COVID-19. J. Artif. Intell. Res. 2020 Nov; 69:807–45. doi: 10.1613/jair.1.12162

[17] Page MJ, et al. The PRISMA 2020 statement: an updated guideline for reporting systematic reviews. BMJ 2021; 372. doi: 10.1136/bmj.n71

[18] Lober, L. Neural networks for dengue forecasting: a systematic review - Data repository. [Accessed 14. Aug. 2026]. 2026 Aug 14. Available from: https://github.com/luizalober/dengue-nn-systematic-review

[19] Tran NT, Xin J, and Zhou G. FWin Transformer for Dengue Prediction Under Climate and Ocean Influence. Machine Learning, Optimization, and Data Science. Cham, Switzerland: Springer, 2025 Mar:160–75. doi: 10.1007/978-3-031-82484-5_12

[20] Andersson VO, Ferreira Birck MA, and Araujo RM. Towards Predicting Dengue Fever Rates Using Convolutional Neural Networks and Street-Level Images. 2018 International Joint Conference on Neural Networks (IJCNN). 2018:1–8. doi: 10.1109/IJCNN.2018.8489567

[21] Andersson VO, Cechinel C, and Araujo RM. Combining Street-level and Aerial Images for Dengue Incidence Rate Estimation. 2019 International Joint Conference on Neural Networks (IJCNN). 2019:1–8. doi: 10.1109/IJCNN.2019.8851749

[22] Livelo ED and Cheng C. Intelligent Dengue Infoveillance Using Gated Recurrent Neural Learning and Cross-Label Frequencies. 2018 IEEE International Conference on Agents (ICA). 2018:2–7. doi: 10.1109/AGENTS.2018.8459963

[23] Araújo JLB, Bomfim R, Filho CINS, Cavalcanti LPG, Neto ASL, Andrade Jr. JS, and Furtado V. The impact of COVID-19 mobility restrictions on dengue transmission in urban areas. PLoS Negl. Trop. Dis. 2024 Nov; 18:e0012644. doi: 10.1371/journal.pntd.0012644

[24] Abdur Rehman N, Saif U, and Chunara R. Deep Landscape Features for Improving Vectorborne Disease Prediction. Proceedings of the IEEE/CVF Conference on Computer Vision and Pattern Recognition (CVPR) Workshops. 2019 Jun.

[25] Dharmawardana KGS, Lokuge JN, Dassanayake PSB, Sirisena ML, Fernando ML, Perera AS, and Lokanathan S. Predictive model for the dengue incidences in Sri Lanka using mobile network big data. 2017 IEEE International Conference on Industrial and Information Systems (ICIIS). Peradeniya: IEEE, 2017 Dec. doi: 10.1109/ICIINFS.2017.8300381

[26] Li Z, Gurgel H, Xu L, Yang L, and Dong J. Improving dengue forecasts by using geospatial big data analysis in Google Earth Engine and the historical dengue information-aided long short term memory modeling. Biology (Basel) 2022 Jan; 11:169. doi: 10.3390/biology11020169

[27] Anggraeni W, Yuniarno EM, Rachmadi RF, Sumpeno S, Pujiadi P, Sugiyanto S, Santoso J, and Purnomo MH. A hybrid EMD-GRNN-PSO in intermittent time-series data for dengue fever forecasting. Expert Syst. Appl. 2024 Mar; 237:121438. doi: 10.1016/j.eswa.2023.121438

[28] Hu J, Horton BP, Yeo TW, Sung JJY, Steve YHL. Mosquito and global dengue cases in a warming world. BMJ Global Health. 2025;10:e014688. 10.1136/bmjgh-2023-014688

[29] Dinh TQ, Le HV, Cao TH, Luong QC, and Diep HT. Forecasting the magnitude of dengue in southern Vietnam. *Intelligent Information and Database Systems*. Lecture Notes in Computer Science. Berlin, Heidelberg: Springer Berlin Heidelberg, 2016:554–63. doi: 10.1007/978-3-662-49381-6_53

[30] Anggraeni W, Wicaksono AA, Yuniarno EM, Rachmadi RF, Sumpeno S, and Purnomo MH. Multilevel analysis of temporal-based spatial factors impact in dengue fever forecasting using RReliefF - deep learning. 2022 IEEE International Conference on Imaging Systems and Techniques (IST). Kaohsiung, Taiwan: IEEE, 2022 Jun. doi: 10.1109/IST55454.2022.9827717

[31] Lu X, Teh SY, Tay CJ, Abu Kassim NF, Fam PS, and Soewono E. Application of multiple linear regression model and long short-term memory with compartmental model to forecast dengue cases in Selangor, Malaysia based on climate variables. Infectious Disease Modelling 2025 Mar; 10:240–56. doi: 10.1016/j.idm.2024.10.007

[32] Dong B, Khan L, Smith M, Trevino J, Zhao B, Hamer GL, Lopez-Lemus UA, Molina AA, Lubinda J, Nguyen USDT, and Haque U. Spatio-temporal dynamics of three diseases caused by *Aedes*-borne arboviruses in Mexico. Commun. Med. (Lond.) 2022 Oct; 2:134. doi: 10.1038/s43856-022-00192-7

[33] Kesorn K, Ongruk P, Chompoosri J, Phumee A, Thavara U, Tawatsin A, and Siriyasatien P. Morbidity rate prediction of dengue hemorrhagic fever (DHF) using the support vector machine and the *Aedes aegypti* infection rate in similar climates and geographical areas. PLoS One 2015 May; 10:e0125049. doi: 10.1371/journal.pone.0125049

[34] Bergero P, Schaposnik LP, and Wang G. Correlations between COVID-19 and dengue obtained via the study of South America, Africa and Southeast Asia during the 2020s. Sci. Rep. 2023 Jan; 13:1–17. doi: 10.1038/s41598-023-27983-9

[35] Roster KO, Martinelli T, Connaughton C, Santillana M, and Rodrigues FA. Impact of the COVID-19 pandemic on dengue in Brazil: Interrupted time series analysis of changes in surveillance and transmission. PLoS Negl. Trop. Dis. 2024 Dec; 18:e0012726. doi: 10.1371/journal.pntd.0012726

[36] Mussumeci E and Codeço Coelho F. Large-scale multivariate forecasting models for Dengue - LSTM versus random forest regression. Spatial and Spatio-temporal Epidemiology 2020; 35:100372. doi: 10.1016/j.sste.2020.100372

[37] Salsabiila NJ and Anggraeni W. CNN+BiGRU-Attention Classification and TiDE-PSO Forecasting Approach for Social Media-based Predictive Analysis of Dengue. International Journal of Intelligent Engineering and Systems 2025.

[38] Anno S, Tsubasa H, Sugita S, Yasumoto S, Lee MA, Sasaki Y, and KO. Challenges and implications of predicting the spatiotemporal distribution of dengue fever outbreak in Chinese Taiwan using remote sensing data and deep learning. Geo-spatial Information Science 2024; 27:1155–61. doi: 10.1080/10095020.2022.2144770

[39] Das D, Maiti S, and Sarma DK. A high-resolution GIS and machine learning approach for targeted disease management and localized risk assessment in an urban setup: A case study from Bhopal, Central India. Acta Trop. 2025 Jul; 267:107662. doi: 10.1016/j.actatropica.2025.107662

[40] Francisco ME, Carvajal TM, and Watanabe K. Hybrid Machine Learning Approach to Zero- Inflated Data Improves Accuracy of Dengue Prediction. PLoS Negl. Trop. Dis. 2024 Oct; 18:e0012599. doi: 10.1371/journal.pntd.0012599

[41] Baquero OS, Santana LMR, and Chiaravalloti-Neto F. Dengue forecasting in São Paulo city with generalized additive models, artificial neural networks and seasonal autoregressive integrated moving average models. PLoS One 2018 Apr; 13:e0195065. doi: 10.1371/journal.pone.0195065

[42] Mustaffa NA, Zahari SM, Farhana NA, Nasir N, and Azil AH. Forecasting the incidence of dengue fever in Malaysia: A comparative analysis of seasonal ARIMA, dynamic harmonic regression, and neural network models. International Journal of Advanced and Applied Sciences 2024 Feb; 11:20–31. doi: 10.21833/ijaas.2024.01.003

[43] Chakraborty T, Chattopadhyay S, and Ghosh I. Forecasting dengue epidemics using a hybrid methodology. Physica A: Statistical Mechanics and its Applications 2019; 527:121266. doi: 10.1016/j.physa.2019.121266

[44] Koh YM, Spindler R, Sandgren M, and JJ. A model comparison algorithm for increased forecast accuracy of dengue fever incidence in Singapore and the auxiliary role of total precipitation information. International Journal of Environmental Health Research 2018; 28:535–52. PMID: 30016117. doi: 10.1080/09603123.2018.1496234

[45] Mahajan A, Sharma N, Aparicio-Obregon S, Alyami H, Alharbi A, Anand D, Sharma M, and Goyal N. A novel stacking-based deterministic ensemble model for infectious disease prediction. Mathematics 2022 May; 10:1714. doi: 10.3390/math10101714

[46] Panja M, Chakraborty T, Nadim SS, Ghosh I, Kumar U, and Liu N. An ensemble neural network approach to forecast Dengue outbreak based on climatic condition. *Chaos*, Solitons & Fractals 2023 Feb; 167:113124. doi: 10.1016/j.chaos.2023.113124

[47] Muhammad F, Hertono GF, and Handari BD. Comparison of activation functions on radial basis function neural network in predicting dengue hemorrhagic fever incidents in DKI Jakarta. AIP Conference Proceedings 2020 Nov; 2296:020017. doi: 10.1063/5.0030453

[48] Hasanah H, Hertono GF, and Sarwinda D. Prediction of dengue incidence in DKI Jakarta using adaptive neuro-fuzzy inference system. International Conference on Science and Applied Science (ICSAS2020). Surakarta, Indonesia: AIP Publishing, 2020. doi: 10.1063/5.0030455

[49] Ronneberger O, Fischer P, and Brox T. U-Net: Convolutional Networks for Biomedical Image Segmentation. *arXiv* 2015 May. doi: 10.48550/arXiv.1505.04597

[50] Ullah MA, Mim AS, Hasan MN, and Sadik MR. Deep Learning Based Forecasting Models of Dengue Outbreak in Bangladesh: Comparative Analysis of LSTM, RNN, and GRU Models Using Multivariate Variables with a Two-Decade Dataset. 2024 International Conference on Smart Systems for Applications in Electrical Sciences (ICSSES). 2024:1–6. doi: 10.1109/ICSSES62373.2024.10561382

[51] Zhao X, Li K, Ang CKE, and Cheong KH. A deep learning based hybrid architecture for weekly dengue incidences forecasting. *Chaos*, Solitons & Fractals 2023 Mar; 168:113170. doi: 10.1016/j.chaos.2023.113170

[52] Ferdousi T, Cohnstaedt LW, and Scoglio CM. A Windowed Correlation-Based Feature Selection Method to Improve Time Series Prediction of Dengue Fever Cases. IEEE Access 2021; 9:141210–22. doi: 10.1109/ACCESS.2021.3120309

[53] García-Garaluz E, Atencia M, Joya G, García-Lagos F, and Sandoval F. Hopfield networks for identification of delay differential equations with an application to dengue fever epidemics in Cuba. Neurocomputing 2011; 74:2691–7. doi: 10.1016/j.neucom.2011.03.022

[54] Li Z. Forecasting Weekly Dengue Cases by Integrating Google Earth Engine-Based Risk Predictor Generation and Google Colab-Based Deep Learning Modeling in Fortaleza and the Federal District, Brazil. International Journal of Environmental Research and Public Health 2022; 19:13555. doi: 10.3390/ijerph192013555

[55] Kakarla SG, Kondeti PK, Vavilala HP, Boddeda GSB, Mopuri R, Kumaraswamy S, Kadiri MR, and Mutheneni SR. Weather integrated multiple machine learning models for prediction of dengue prevalence in India. Int. J. Biometeorol. 2023 Feb; 67:285–97. doi: 10.1007/s00484-022-02405-z

[56] Chen X and Moraga P. Forecasting dengue across Brazil with LSTM neural networks and SHAP-driven lagged climate and spatial effects. BMC Public Health 2025 Dec; 25:1–22. doi: 10.1186/s12889-025-22106-7

[57] Ceballos-Arroyo, et al. Towards a machine learning-based approach to forecasting Dengue virus outbreaks in Colombian cities: a case study: Medellín, Antioquia. 15th International Symposium on Medical Information Processing and Analysis. Ed. by Brieva J, Romero E, and Lepore N. Medellín, Colombia: SPIE, 2020 Jan. doi: 10.1117/12.2547001

[58] Anggraeni W et al. Artificial Neural Network for Health Data Forecasting, Case Study: Number of Dengue Hemorrhagic Fever Cases in Malang Regency, Indonesia. 2018 International Conference on Electrical Engineering and Computer Science (ICECOS). 2018, 207–12. doi: 10.1109/ICECOS.2018.8605254

[59] Sukama Y, Hertono GF, Handari BD, and Aldila D. Comparing activation functions in predicting dengue hemorrhagic fever cases in DKI Jakarta using recurrent neural networks. AIP Conference Proceedings 2020 Nov; 2296:020059. doi: 10.1063/5.0030456

[60] Herath PHMN, Perera AAI, and HW. Prediction of Dengue Outbreaks in Sri Lanka using Artificial Neural Networks. International Journal of Computer Applications 2014 Sep; 101:1–5. doi: 10.5120/17760-8862

[61] Rachata N, Charoenkwan P, Yooyativong T, Chamnongthal K, Lursinsap C, and Higuchi K. Automatic Prediction System of Dengue Haemorrhagic-Fever Outbreak Risk by Using Entropy and Artificial Neural Network. 2008 International Symposium on Communications and Information Technologies. 2008:210–4. doi: 10.1109/ISCIT.2008.4700184

[62] Dhaked DK, Sharma O, Gopal Y, and Gopal R. Prediction of dengue patients using deep learning methods amid complex weather conditions in Jaipur, India. Discover Public Health 2025 Dec; 22:1–13. doi: 10.1186/s12982-025-00448-2

[63] Jaya IGNM, Andriyana Y, Tantular B, Pangastuti SS, and Kristiani F. Spatiotemporal Dengue Forecasting for Sustainable Public Health in Bandung, Indonesia: A Comparative Study of Classical, Machine Learning, and Bayesian Models. Sustainability 2025 Jul; 17:6777. doi: 10.3390/su17156777

[64] Akter T, Islam MT, Hossain MF, and Ullah MS. A Comparative Study between Time Series and Machine Learning Technique to Predict Dengue Fever in Dhaka City. Discrete Dynamics in Nature and Society 2024; 2024:2757381. doi: 10.1155/2024/2757381

[65] Abeyrathna KD, Granmo OC, Zhang X, and Goodwin M. A Scheme for Continuous Input to the Tsetlin Machine with Applications to Forecasting Disease Outbreaks. Advances and Trends in Artificial Intelligence. From Theory to Practice. Ed. by Wotawa F, Friedrich G, Pill I, Koitz-Hristov R, and Ali M. Cham: Springer International Publishing, 2019:564–78. doi: 10.1007/978-3-030-22999-3_49

[66] Xu J, Xu K, Li Z, Meng F, Tu T, Xu L, and Liu Q. Forecast of Dengue Cases in 20 Chinese Cities Based on the Deep Learning Method. Int. J. Environ. Res. Public Health 2020 Jan; 17:453. doi: 10.3390/ijerph17020453

[67] Sérgio de Souza L, et al. Forecast of Dengue Cases based on the Deep Learning Approach: A Case Study for a Brazilian City. Proceedings of the 3rd International Conference on Deep Learning Theory and Applications - DeLTA. INSTICC. SciTePress, 2022:71–6. doi: 10.5220/0011135500003277

[68] Sousa LB, Von Zuben CJ, and Von Zuben FJ. Regularized linear and nonlinear autoregressive models for dengue confirmed-cases prediction. Unconventional Computation and Natural Computation. Lecture Notes in Computer Science. Cham: Springer International Publishing, 2015:121–31. doi: 10.1007/978-3-319-21819-9_8

[69] Sekmen A and Bilgin B. Manifold-based approach for neural network robustness analysis. Commun. Eng. 2024 Aug; 3:1–13. doi: 10.1038/s44172-024-00263-8

[70] Linardatos P, Papastefanopoulos V, and Kotsiantis S. Explainable AI: A Review of Machine Learning Interpretability Methods. Entropy 2020 Dec; 23:18. doi: 10.3390/e23010018

[71] Messina JP et al. The current and future global distribution and population at risk of dengue. Nat. Microbiol. 2019 Sep; 4:1508–15. doi: 10.1038/s41564-019-0476-8

[72] Ganem F, et al. Mosqlimate: a platform to providing automatable access to data and forecasting models for arbovirus disease. 2024. arXiv: 2410.18945

[73] Necesito I et al. Combination of univariate long-short term memory network and wavelet transform for predicting dengue case density in the National Capital Region, the Philippines. The Southeast Asian Journal of Tropical Medicine and Public Health 2021 Jul; 52:479–94.

[74] Nguyen VH et al. Deep learning models for forecasting dengue fever based on climate data in Vietnam. PLoS Negl. Trop. Dis. 2022 Jun; 16:e0010509. doi: 10.1371/journal.pntd.0010509

[75] Rahayu I, Nurhasanah N, and Adriat R. Prediction of Dengue Hemorrhagic Fever Cases Based on Weather Parameters Using Back Propagation Neural Networks (Case Study in Pontianak City). Jurnal Pendidikan Fisika Indonesia 2019; 15:114–21. doi: 10.15294/jpfi.v15i2.19633

[76] Bogado JV, Stalder DH, Schaerer CE, and Gómez-Guerrero S. Time Series Clustering to Improve Dengue Cases Forecasting with Deep Learning. 2021 XLVII Latin American Computing Conference (CLEI). 2021:1–10. doi: 10.1109/CLEI53233.2021.9640130

[77] Soemsap T, Wongthanavasu S, and Satimai W. Forecasting number of dengue patients using cellular automata model. 2014 International Electrical Engineering Congress (iEECON). 2014:1–4. doi: 10.1109/iEECON.2014.6925876

[78] Conde-Gutiérrez RA, Colorado D, Márquez-Nolasco A, and Gonzalez-Flores PB. Parallel prediction of dengue cases with different risks in Mexico using an artificial neural network model considering meteorological data. Int. J. Biometeorol. 2024 Jun; 68:1043–60. doi: 10.1007/s00484-024-02643-3

[79] Anno S et al. Spatiotemporal dengue fever hotspots associated with climatic factors in Taiwan including outbreak predictions based on machine-learning. Geospat. Health 2019 Nov; 14. doi: 10.4081/gh.2019.771

[80] Navarro Valencia V et al. Assessing the effect of climate variables on the incidence of dengue cases in the Metropolitan Region of Panama City. Int. J. Environ. Res. Public Health 2021 Nov; 18:12108. doi: 10.3390/ijerph182212108

[81] Aburas HM, Gultekin Cetiner B, and Sari M. Dengue confirmed-cases prediction: A neural network model. Expert Systems with Applications 2010; 37:4256–60. doi: 10.1016/j.eswa.2011.04.120

[82] Rajendran NM et al. Communicable Disease Prediction Using Machine Learning and Deep Learning Algorithms. Inventive Communication and Computational Technologies. Ed. by Ranganathan G, Papakostas GA, and Rocha Á. Singapore: Springer Nature Singapore, 2023:979–92. doi: 10.1007/978-981-99-5166-6_66

[83] Datoc HI, Caparas R, and Caro J. Forecasting and data visualization of dengue spread in the Philippine Visayas island group. 2016 7th International Conference on Information, Intelligence, Systems & Applications (IISA). Chalkidiki, Greece: IEEE, 2016 Jul. doi: 10.1109/IISA.2016.7785420

[84] Yusof Y and Mustaffa Z. Dengue outbreak prediction: A least squares support vector machines approach. Int. J. Comput. Theory Eng. 2011:489–93. doi: 10.7763/IJCTE.2011.V3.355

[85] Isidoro C, Fachada N, Barata F, and Rosa A. Agent-Based Model of *Aedes aegypti* Population Dynamics. *Progress in Artificial Intelligence*. Berlin, Germany: Springer, 2009:53–64. doi: 10.1007/978-3-642-04686-5_5

[86] Johansson MA, Reich NG, Hota A, Brownstein JS, and Santillana M. Evaluating the performance of infectious disease forecasts: A comparison of climate-driven and seasonal dengue forecasts for Mexico. Sci. Rep. 2016 Sep; 6:1–11. doi: 10.1038/srep33707

[87] Riley P, Ben-Nun M, Turtle J, Bacon D, and Riley S. SARIMA Forecasts of Dengue Incidence in Brazil, Mexico, Singapore, Sri Lanka, and Thailand: Model Performance and the Significance of Reporting Delays. medRxiv 2020 Jun: 2020.06.26.20141093. doi: 10.1101/2020.06.26.20141093

[88] Anwar SM, Majid M, Qayyum A, Awais M, Alnowami M, and Khan MK. Medical Image Analysis using Convolutional Neural Networks: A Review. J. Med. Syst. 2018 Nov; 42:1–13. doi: 10.1007/s10916-018-1088-1

[89] Papik K, Molnar B, Schaefer R, Dombovari Z, Tulassay Z, and Feher J. Application of neural networks in medicine - a review. Diagnostics and Medical Technology 1998 May; 4:MT538–MT546.

[90] Abiodun OI, Jantan A, Omolara AE, Dada KV, Mohamed NA, and Arshad H. State-of- the-art in artificial neural network applications: A survey. Heliyon 2018 Nov; 4:e00938. doi: 10.1016/j.heliyon.2018.e00938

[91] Mienye ID, Swart TG, and Obaido G. Recurrent Neural Networks: A Comprehensive Review of Architectures, Variants, and Applications. Information 2024 Aug; 15:517. doi: 10.3390/info15090517

[92] Cong S and Zhou Y. A review of convolutional neural network architectures and their optimizations. Artif. Intell. Rev. 2023 Mar;56:1905–69. doi: 10.1007/s10462-022-10213-5

[93] Rosenblatt F. The perceptron: a probabilistic model for information storage and organization in the brain. Psychol. Rev. 1958 Nov; 65:386–408. doi: 10.1037/h0042519

[94] Chowdhury AH. Comparison of Deep Learning and Gradient Boosting: ANN Versus XG- Boost for Climate-Based Dengue Prediction in Bangladesh. Health Science Reports 2025 Apr; 8:e70714. doi: 10.1002/hsr2.70714

[95] Zahiruddin H, Zukarnain ZA, and Wijaya A. A scoping review and bibliometric analysis (ScoRBA) on dengue infection and machine learning research. JOIV: International Journal on Informatics Visualization 2024 Dec; 8:2203–10. doi: 10.62527/joiv.8.4.2249

